# Cholinergic innervation topography in GBA-associated *de novo* Parkinson’s disease patients

**DOI:** 10.1101/2023.06.16.23290981

**Authors:** Sofie Slingerland, Sygrid van der Zee, Giulia Carli, Anne C. Slomp, Jeffrey M. Boertien, Nicolaas I. Bohnen, Roger L. Albin, Teus van Laar

## Abstract

The most common genetic risk factors for Parkinson’s disease are *GBA1* mutations, encoding the lysosomal enzyme glucocerebrosidase. Patients with *GBA1* mutations (GBA-PD) exhibit earlier age of onset and faster disease progression with more severe cognitive impairments, postural instability, and gait problems. The GBA-PD features suggests more severe cholinergic system pathologies. PET imaging with the vesicular acetylcholine transporter ligand [^18^F]-F-fluoroethoxybenzovesamicol ([^18^F]FEOBV PET) provides the opportunity to investigate cholinergic systems changes and their relationship to clinical features of GBA-PD.

One hundred and twenty three newly diagnosed, treatment-naive Parkinson’s disease subjects – with confirmed presynaptic dopaminergic deficits on PET imaging – were included, all part of the Dutch Parkinson Cohort (DUPARC) study. Full *GBA1* sequencing of saliva samples was performed to evaluate *GBA1* variants. Patients underwent extensive neuropsychological assessment assessing all cognitive domains, motor evaluation with the Unified Parkinson’s disease Rating Scale, brain MRI, dopaminergic PET to measure striatal-to-occipital ratios of the putamen and [^18^F]FEOBV PET. We investigated differences in regional cholinergic innervation between GBA-PD carriers and non-*GBA1* mutation carriers (non-GBA-PD), using voxel-wise and volume-of-interest (VOI)-based approaches. The degree of overlap between t-maps from two-sample t-test models was quantified using the Dice similarity coefficient.

Seventeen (13.8%) subjects had a *GBA1* mutation. No significant differences were found in the clinical features and dopaminergic ratios between GBA-PD and non-GBA-PD at diagnosis. Lower [^18^F]FEOBV binding was found in both the GBA-PD and non-GBA-PD group compared to controls. Dice (*P* < 0.05, cluster size 100) showed a good overlap (0.6233) between the GBA-PD and non-GBA-PD maps. GBA-PD patients showed more widespread reduction in [^18^F]FEOBV binding than non-GBA-PD when compared to controls in occipital, parietal, temporal, and frontal cortices (*P* < 0.05, FDR-corrected). In VOI analyses (Bonferroni corrected), the left cuneus, entorhinal cortex, fusiform gyrus and supramarginal gyrus were more affected in GBA-PD.

*De novo* GBA-PD show a distinct topography of regional cholinergic terminal ligand binding, including both higher and lower binding regions. Although the Parkinson’s disease groups were not distinguishable clinically, in comparison to healthy controls, GBA-PD showed more extensive cholinergic denervation compared to non-GBA-PD. Our results suggest that *de novo* GBA-PD and non-GBA-PD show differential patterns of cholinergic system changes before clinical differences are observable.

## Introduction

The most common genetic risk factors for Parkinson’s disease are heterozygous mutations of *GBA1*, encoding the lysosomal enzyme glucocerebrosidase.^1, 2^ Parkinson’s disease patients with *GBA1* mutations (GBA-PD) exhibit more aggressive disease than Parkinson’s disease patients without *GBA1* mutation (non-GBA-PD).^3, 4^ GBA-PD is associated with earlier and faster cognitive declines.^5^ In GBA-PD, risk of dementia is increased up to 6-fold and starts approximately 5 years earlier when compared to unselected Parkinson’s disease populations.^6–9^ Neuropsychiatric symptoms are more frequently observed in GBA-PD,^10, 11^ and GBA-PD is associated with the postural instability and gait difficulties (PIGD) motor subtype.^5, 12^

Postural instability and gait problems are associated with brain impaired cholinergic systems, independent of dopaminergic deficits.^13^ Cognitive declines and cholinergic dysfunctions are interrelated.^14, 15^ Previous *in vivo* Parkinson’s disease neuroimaging assessments, using acetylcholinesterase PET, demonstrated regional cholinergic synapses deficits primarily in posterior cortices.^16^ Cholinergic denervation can already be present in early Parkinson’s disease and is more widespread and severe in Parkinson’s disease dementia.^17^ We demonstrated previously that recently diagnosed, treatment-naive Parkinson’s disease patients, relative to controls, show bidirectional changes in a specific marker of presynaptic cholinergic terminals. Increased cholinergic terminal ligand binding in early Parkinson’s disease, primarily in cognitively intact subjects, suggests compensatory cholinergic upregulation in this group.^18^

GBA-PD patients often show faster progression of both motor and nonmotor features with greater risks of cognitive impairments and postural instability-gait disorders, both features associated with cholinergic deficits.^4^ Analysis of cholinergic system changes in GBA-PD is needed to understand the pathophysiology of these morbid Parkinson’s disease features. There is little data on brain cholinergic systems in GBA-PD. In-depth characterisation of GBA-PD is important to validate GBA-PD as a potentially distinct subtype for potential personalised treatment approaches. More detailed phenotyping of early GBA-PD is likely to provide useful prognostic information.

This study investigated the clinical phenotype and regional density of cholinergic terminals in newly diagnosed, treatment-naive Parkinson’s disease patients comparing, GBA-PD with non-GBA-PD and healthy controls (HC) with [^18^F]Fluoroethoxybenzovesamicol ([^18^F]FEOBV) PET. [^18^F]FEOBV binds to the vesicular acetylcholine transporter and is a valid marker of cholinergic terminal integrity in both healthy and diseased brains.^19–21^

## Materials and methods

### Participants

One hundred and twenty three newly diagnosed, treatment-naive Parkinson’s disease patients and 10 healthy controls (HC) were included in the Dutch Parkinson Cohort (DUPARC) study.^22^ Inclusion criteria for patients consisted of Clinically Probable Parkinson’s disease diagnosis by a movement disorders specialist according to Movement Disorders Society (MDS) Clinical Diagnosis Criteria for Parkinson’s disease,^23^ and with confirmed dopaminergic striatal deficits on [^18^F]-3,4-dihydroxy-6-18F-fluoro-I-phenylalanine ([^18^F]FDOPA) PET. HCs had a normal neurological examination and no histories of neurological or psychiatric disorders. Exclusion criteria included inability to provide written informed consent, use of dopaminergic and/or (anti-)cholinergic medication and estimated low premorbid intelligence level (estimated IQ <70; on the Dutch Adult Reading test).^24^ The study was conducted according to the Good Clinical Practice guidelines and the Declaration of Helsinki. All subjects gave written informed consent, and the local ethics committee approved the study.

### Genotyping

Saliva was obtained from Parkinson’s disease patients using Oragene DNA OG-500 tubes (DNA Genotek). DNA isolation, next generation sequencing (NGS), and data analysis were performed by GenomeScan B.V., Leiden, the Netherlands. Primers were selected to unambiguously sequence the functional *GBA1*-gene and not the pseudogene, using long-range PCR. Fragments were amplified using PCR and sequenced using Illumina cBot and HiSeq 400 as described previously.^25^ The allelic nomenclature of the *GBA1* variants is given.

### Matching procedure

Genotyping led to unbalanced groups in terms of sample size (see *Results and Table 2*), where non-GBA-PD patients represented the majority of participants, consistent with the known epidemiology of GBA-PD mutations.^25^ Since equal-sized groups maximise statistical power,^26^ we created non-GBA-PD sub-groups with identical subject numbers as the GBA-PD group. Non-GBA-PD patients were matched 1:1 for age and gender with GBA-PD subjects by case-control matching (SPSS version 28). Match tolerances were zero for gender and 2 years for age (non-GBA-PD group 1). A second matched non-GBA-PD group was created to validate the results using a similar procedure after excluding the first already matched non-GBA-PD subjects. The match tolerances were zero for gender and 6 years for age in the second non-GBA-PD group (non-GBA-PD group 2).

### Clinical examination

All Parkinson’s disease participants underwent comprehensive neuropsychological assessment covering the main cognitive domains: memory, attention, executive function, language and visuospatial abilities. A selection of outcome measures of tests and subtests of the cognitive test battery was made a priori, allowing level II criteria for Parkinson’s disease with mild cognitive impairment (PD-MCI; *Table 1*). Below-threshold performance on at least two neuropsychological tests was required for PD-MCI classification.^27, 28^. Scores >1.5 standard deviations (SD) below normative values were considered abnormal. Patients were categorised as either Parkinson’s disease with normal cognition or PD-MCI. All (sub)test scores were transformed into standardised z-scores based on established test-specific normative data. Test scores for the Boston naming test and the Location learning test were transformed into standardised z-scores based on a sample of 108 HC (58 males, ages ranging between 41 and 84 years; M = 64.49 years, SD = 8.08 years) collected as part of the DUPARC study. All z-scores within one cognitive domain were averaged to define domain-specific z-score for each of the five domains. HC subjects undergoing [^18^F]FEOBV PET imaging underwent cognitive testing using the Montreal Cognitive Assessment test. Visual difficulties, color blindness, speech problems, and significant mood disorders that possibly influenced performance on the neuropsychological assessment and MCI grouping were taken in to account prior to data analysis, and if necessary excluded.

**Table 1:** Cognitive scales applied per domain.

| Domain | Cognitive scale |
| --- | --- |
| Memory | RAVLT - Immediate recall |
|  | RAVLT - Delayed recall |
|  | Location learning test |
| Attention | Trial-making test - A |
|  | Stroop - I |
| Visuospatial | Judgement of line orientation |
| Executive | Trial-making test - B |
|  | Stroop - III |
|  | Letter-fluency |
| Language | Boston naming test |
|  | Semantic fluency - occupation |
|  | Semantic fluency - animals |
| Abbreviations: RAVLT - Rey Auditory Verbal Learning Test. |  |

Clinical motor performance was examined using the MDS-revised Unified Parkinson’s Disease Rating Scale part III (UDPRS-III). Specific items from the UPDRS parts II and III were used for classification of motor phenotype, using standard criteri*a*.^29^ Participants were classified as tremor dominant, postural instability and gait difficulty, and indeterminate motor phenotypes. Additional clinical assessments included the Hospital Anxiety and Depression Scale, Non-Motor Symptoms Questionnaire, NMS-REM Sleep Behavior Disorder Screening Questionnaire, and Burghart Sniffin’ Sticks 12 Tests.^22^

### Image acquisition

All subjects underwent brain MRI, [^18^F]FDOPA PET and VAChT PET imaging with [^18^F]FEOBV. MRI imaging of Parkinson’s disease subjects was acquired using Siemens Magnetom Prisma 3-Tesla magnetic resonance imaging scanners (Best, Netherlands), equipped with SENSE-8 channel head coil. For each subject, anatomical T1-weighted images were obtained using a sagittal 3-dimensional gradient-echo T1 weighted sequence with 0.9×0.9×0.9mm acquisition. HC subjects underwent a T1-weighted MRI scan (3T Intera, Philips, The Netherlands) with 1.0 x 1.0 x 1.0 mm acquisition. [^18^F]FEOBV imaging was performed on the same day as MR imaging. For [^18^F]FEOBV PET imaging, participants first underwent lose-dose computed tomography (CT) for attenuation and scatter correction using either a Biograph 40-mCT or 64-mCT (Siemens Healthcare, USA). Both scanners were EARL certified, had the same software version, and used identical acquisition and reconstruction protocols and PET detectors. Thirty minute scans (in six 5-minute frames) were acquired at 210 minutes post-bolus injection of [^18^F]FEOBV. [^18^F]FDOPA PET was performed after at least six hours of fasting (four hours for diabetic patients). Participants were premedicated with carbidopa 60 minutes before receiving 200MBq of the FDOPA tracer, 90 minutes after which the PET-scan was formed on a Siemens HR+ camera.

### Image pre-processing and analysis

We visually checked all images and set the origin on the anterior commissure. Statistical parametric mapping (SPM12; Wellcome Trust Centre for Neuroimaging, London, UK) software was used to realign the PET imaging frames (six 5-minute frames) within subjects to reduce the effects of subject motion during the imaging session. We co-registered [^18^F]FEOBV PET images to their corresponding T1-weighted volumetric scan (MRI) in native space. We then computed parametric images to reflect the Distribution Volume Ratios (DVR) of [^18^F]FEOBV in the brain using supratentorial white matter above the ventricles as the reference region.^30^ PMOD version 3.8 was used to quantify [^18^F]FDOPA striatal-to-occipital ratios of the putamen.^31^

#### Volume of interest (VOIs) analyses

Each MRI T1-weighted image underwent FreeSufer-based segmentation using the recon-all pipeline with the default settings. Cortical and subcortical labels from Deskian/Killiany and Aseg atlases parcellation were used to identify grey matter subject-specific VOIs (VOI list; *Supplementary Table 1*). We extracted the mean DVR values from [^18^F]FEOBV PET parametric images in native space. We use MRI-derived VOIs, and parametric PET images registered to MRI in native space, to mitigate partial volume effects and not introduce spill-over artifacts due to image translation from native to Montreal Neurological Institute spaces.

#### Voxel-wise analyses

A Muller-Gartner partial-volume correction method was used to remove partial volume effects in the parametric PET images. Then, partial volume effect corrected parametric images were normalised to a study-specific template in Montreal Neurological Institute space using high-dimensional DARTEL registration. The study-specific Montreal Neurological Institute template was obtained via the DARTEL normalisation procedure on MRI T1-weighted images. Last, we applied an 8 mm full width at half maximum smoothing filter to improve the signal-to-noise ratio.

After the above-described pre-processing, we assessed differences in [^18^F]FEOBV binding. First, we evaluated cholinergic topography in both GBA-PD and non-GBA-PD groups, comparing them to the topography of cholinergic terminals in healthy controls. Second, we directly compared GBA-PD and non-GBA-PD. This analysis was repeated with the second non-GBA-PD group for validation.

#### GBA-PD and non-GBA-PD *vs.* HCs

We wanted to investigate the differences between HC and patients’ groups to identify the lower VAChT binding (DVR significantly lower than HC) and higher VAChT binding (DVR significantly higher than HC) in each Parkinson’s disease group. We implemented two-sample t-test models in SPM12 for the following voxel-wise comparisons: i) GBA-PD *vs.* HC and ii) non-GBA-PD group 1 *vs*. HC. We entered age as a covariate of no interest in all comparisons to control age differences (*Table 2*). The threshold was set at *P* < 0.05, FDR-corrected at the voxel level. We used a cluster size of 100 in all analyses. We then compared the topographical similarity of the voxel-wise t-maps resulting from these comparisons (non-GBA-PD group 1 *vs*. HC and GBA-PD *vs*. HC).

**Table 2.**
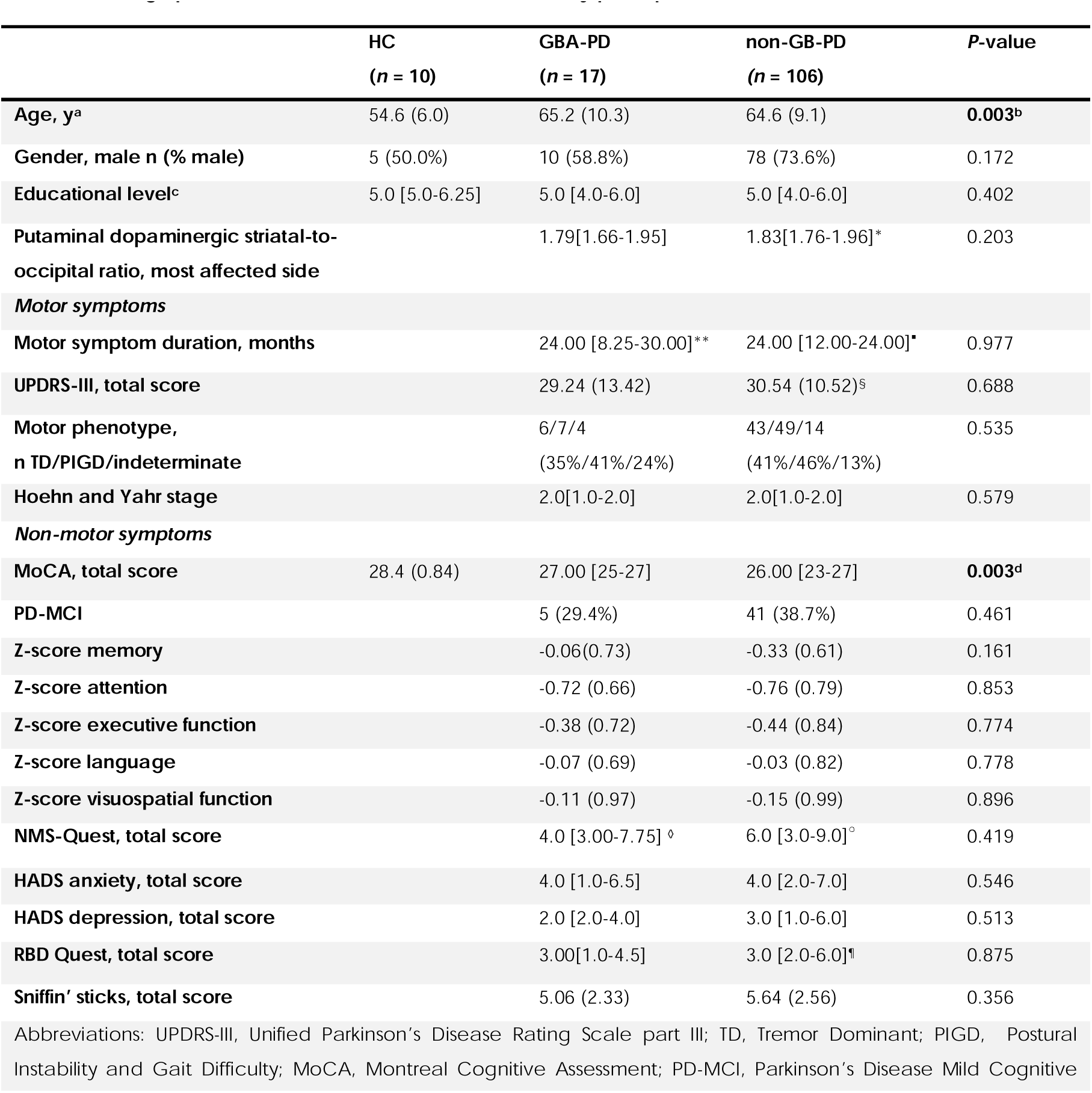
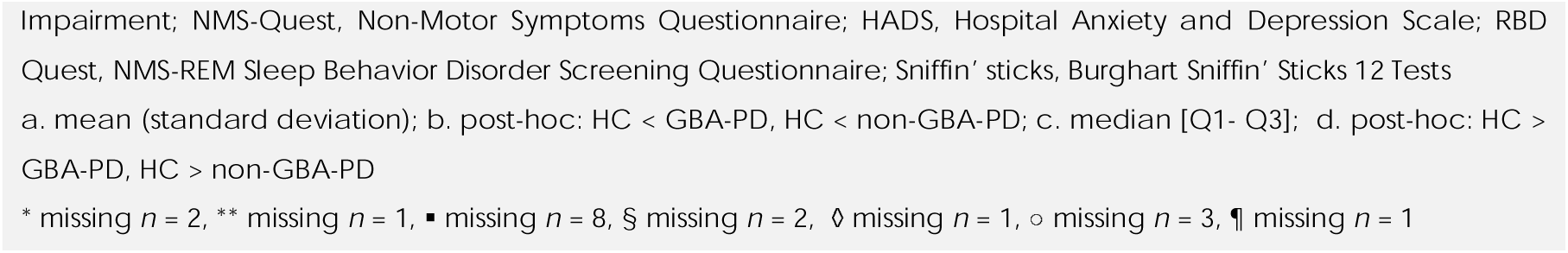
Demographics and Clinical Characteristics of the study participants.

We used the Dice similarity coefficient,^32, 33^ to quantify the degree of overlaps between t-maps resulting from each two-sample t-test SPM model: t-maps of lower VAChT binding – i.e. the brain clusters significantly lower than HC – and t-maps of higher VAChT binding – i.e. the clusters significantly higher than HC. This metric measures volume overlaps between 2 regions divided by their mean volume. It is interpreted as follows: < 0.4, poor; 0.2-0.4, fair; 0.4-0.6, moderate; 0.6-0.8, good; and > 0.8, excellent agreement.

#### GBA *vs.* non-GBA-PD comparisons

We implemented two-sample t-test models in SPM12 for three different comparisons: i) GBA-PD *vs.* non-GBA-PD whole group, ii) GBA-PD *vs.* non-GBA-PD group 1 and iii) GBA-PD *vs.* non-GBA-PD group 2. The threshold was set at *P* < 0.05, FDR-corrected at the voxel level.

### Statistical analysis

We evaluated the differences in demographic, clinical, and cognitive variables and cholinergic VOIs DVR values among HC, GBA-PD and non-GBA-PD groups using ANOVA for continuous variables and χ^2^ testing for dichotomous variables. HCs were significantly younger than the PD groups and we used age as a covariate in the voxel-based and VOI analyses comparing Parkinson’s disease groups and HCs. We used a Multivariate Generalized Linear Model to compare mean DVR values from the extracted VOIs among clinical groups (GBA-PD, non-GBA-PD group 1, and HC); age was entered as covariate of no interest. The statistical threshold was set at *P* < 0.05; Bonferroni correction was used for multiple comparisons. Direct comparisons between the GBA and non-GBA-PD groups were performed using an independent sample t-test or a Mann-Whitney U test, for normally and non-normally distributed data, respectively. The striatal-to-occipital ratios of the [^18^F]FDOPA uptake were quantified by dividing the mean activity of the putamen as the volume-of-interest by the mean occipital value. Statistical analyses were performed using SPSS Statistics for Windows, Version 28.0 (IBM Statistics, USA).

### Data availability

The data that support the findings of this study are available from the corresponding author, upon reasonable request.

## Results

### *GBA1* genetic screening

In total, 17 of the 123 (13.8%) subjects in our DUPARC cohort had a GBA nonsynonymous variant. Seven different mutations were identified. The E326K (6 subjects), T369M (5 subjects) and E326K / D140H (2 subjects) mutations were the most frequently occurring variants. All *GBA1* exonic and splice-site variants are listed in the supplementary materials.

### Demographic and clinical evaluation

The 123 *de novo* PD patients (88 males) had a mean (SD) age of 64.7 (9.2) years and a mean (SD) UPDRS-III score of 30.5 (11.0). Based on MDS PD-MCI level II criteria, 46 (37.4%) of PD patients were classified as PD-MCI: 5 out of 17 (29.4%) GBA-PD and 41 out of 123 (38.7%) non-GBA-PD. HC subjects had a significantly lower age and higher Montreal Cognitive Assessment score than both Parkinson’s disease groups. Comparisons between GBA-PD and non-GBA-PD groups showed no significant difference in demographic, clinical, and cognitive variables (*Table 2*). Also, putaminal dopaminergic striatal-to-occipital ratios of the most affected side revealed no significant difference between the Parkinson’s disease groups (*Table 2*). Similarly, no differences emerged comparing GBA-PD and the matched non-GBA-PD groups (*Supplementary Tables 3 and 4*).

### Cholinergic voxel-based comparisons

#### GBA-PD and non-GBA-PD group *vs.* HC

##### Lower [^18^F]FEOBV binding

Lower [^18^F]FEOBV binding was found in both GBA-PD and non-GBA-PD group when compared to HCs. GBA-PD exhibited widespread lower [^18^F]FEOBV binding involving the occipital, parietal, temporal, and superior and anterior frontal cortices (*Fig. 1A; P < 0.05, FDR-corrected*). Non-GBA-PD subjects exhibited lower [^18^F]FEOBV binding with significant clusters in bilateral inferior and middle occipital gyri, inferior parietal gyri (L > R) and insula (*Fig. 1B; P < 0.05, FDR-corrected*). The Dice similarity coefficient (*P* < 0.05, cluster size 100) showed spatial overlap of 0.6233 (good) between the two maps (GBA-PD *vs.* HC and non-GB-PD *vs.* HC). This Dice coefficient indicates that the GBA-PD and non-GBA-PD group exhibit overlap in the distribution of diminished [^18^F]FEOBV binding, but also some differences in the topography of [^18^F]FEOBV binding deficits.

**Figure 1:**
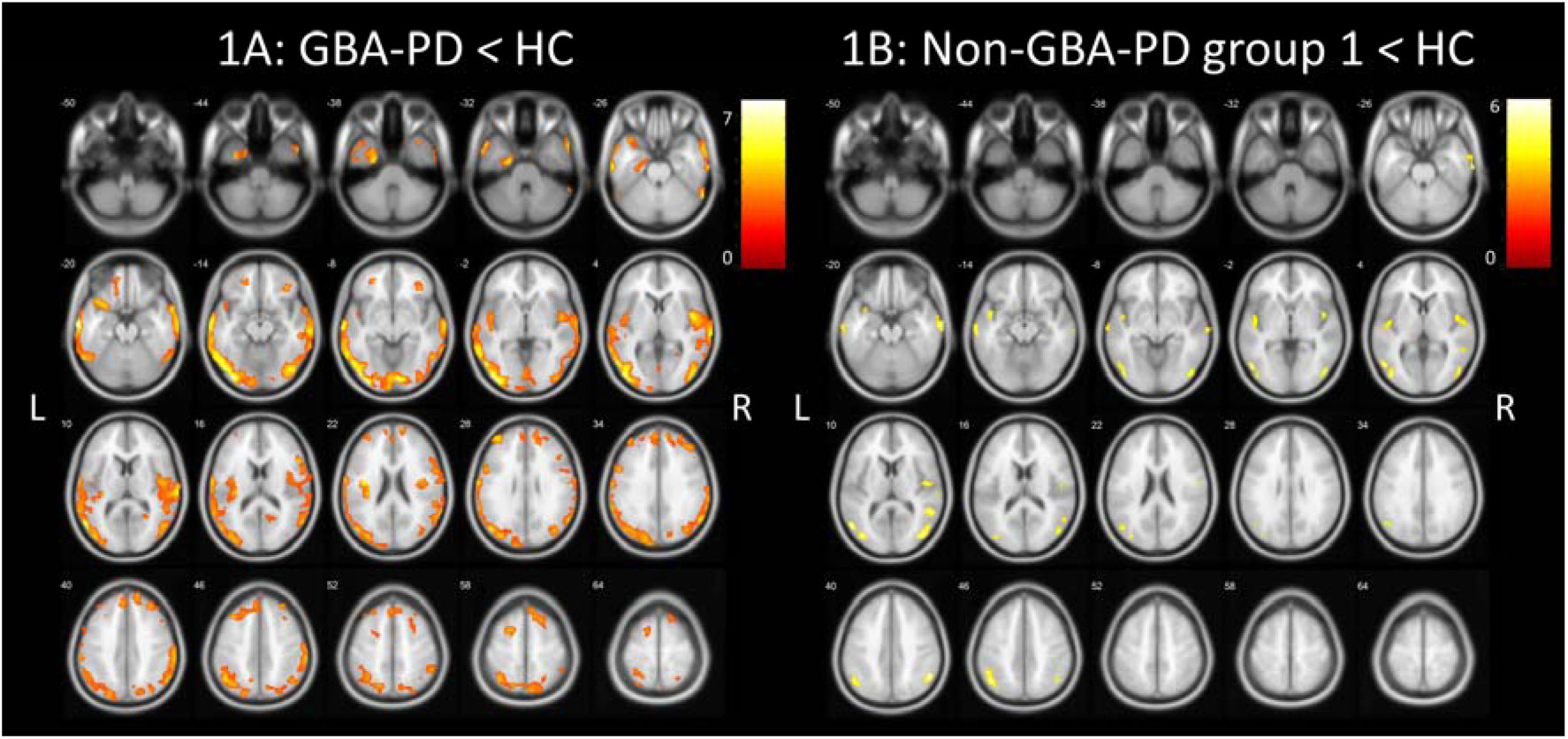
Whole brain voxel-based analyses showing significantly lower VAChT binding (*P* < 0.05, FDR-corrected at voxel-level, cluster size 100) in GBA-PD, *n* = 17 (2A) and non-GBA-PD, *n* = 17 (2B) compared to HC, controlled for age. L, left; R, right.

##### Higher [^18^F]FEOBV binding

FDR corrected data showed no significant voxels with higher [^18^F]FEOBV binding when GBA-PD was compared to HCs. Higher [^18^F]FEOBV binding emerged in non-GBA-PD in the left and right cerebellum, vermis, left thalamus, and the left more than right precentral gyrus compared to controls (*Fig. 2; P < 0.05, FDR corrected*). An additional exploratory FDR-uncorrected analysis was performed with a *P* < 0.001 (see *Fig 3.).* GBA-PD showed higher [^18^F]FEOBV binding than HC in both thalami, left putamen and precentral gyrus and right medial frontal gyrus (Brodmann area 25) and right gyrus rectus (*Fig. 3A; P < 0.001, uncorrected*). In contrast to the GBA-PD group, higher [^18^F]FEOBV was located at the cerebellum in non-GBA-PD (*Fig. 3B; P < 0.001, uncorrected*). Dice results showed a poor overlap (0.0943) when using a *P* < 0.001 and a cluster size of 100 between both binary maps.

**Figure 2:**
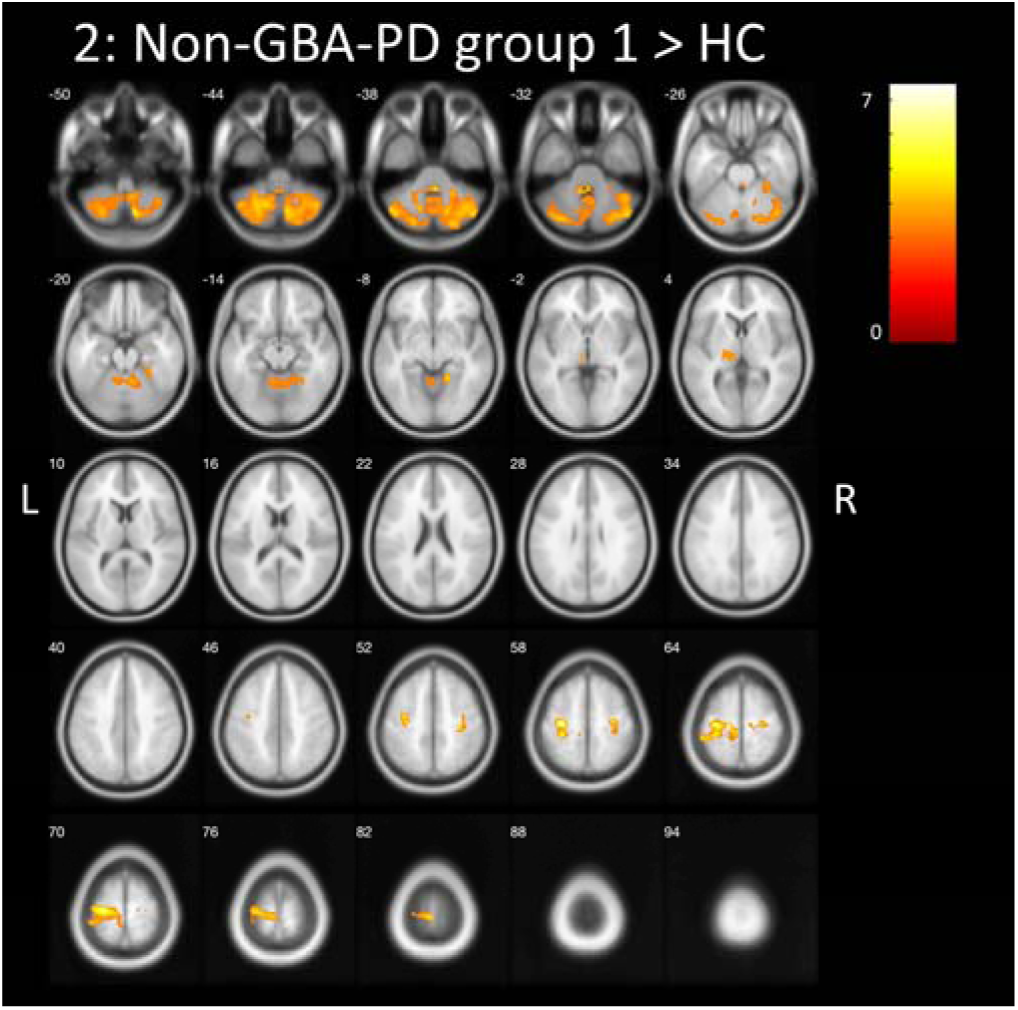
Whole brain voxel-based analyses showing significant higher VAChT binding (*P* < 0.05, FDR-corrected at voxel-level, cluster size 100) in non-GBA-PD, *n* = 17 compared to HC, *n* = 10, controlled for age. L, left; R, right. No voxel survived significance in the GBA-PD *vs.* HC comparison and is therefore not shown.

**Figure 3:**
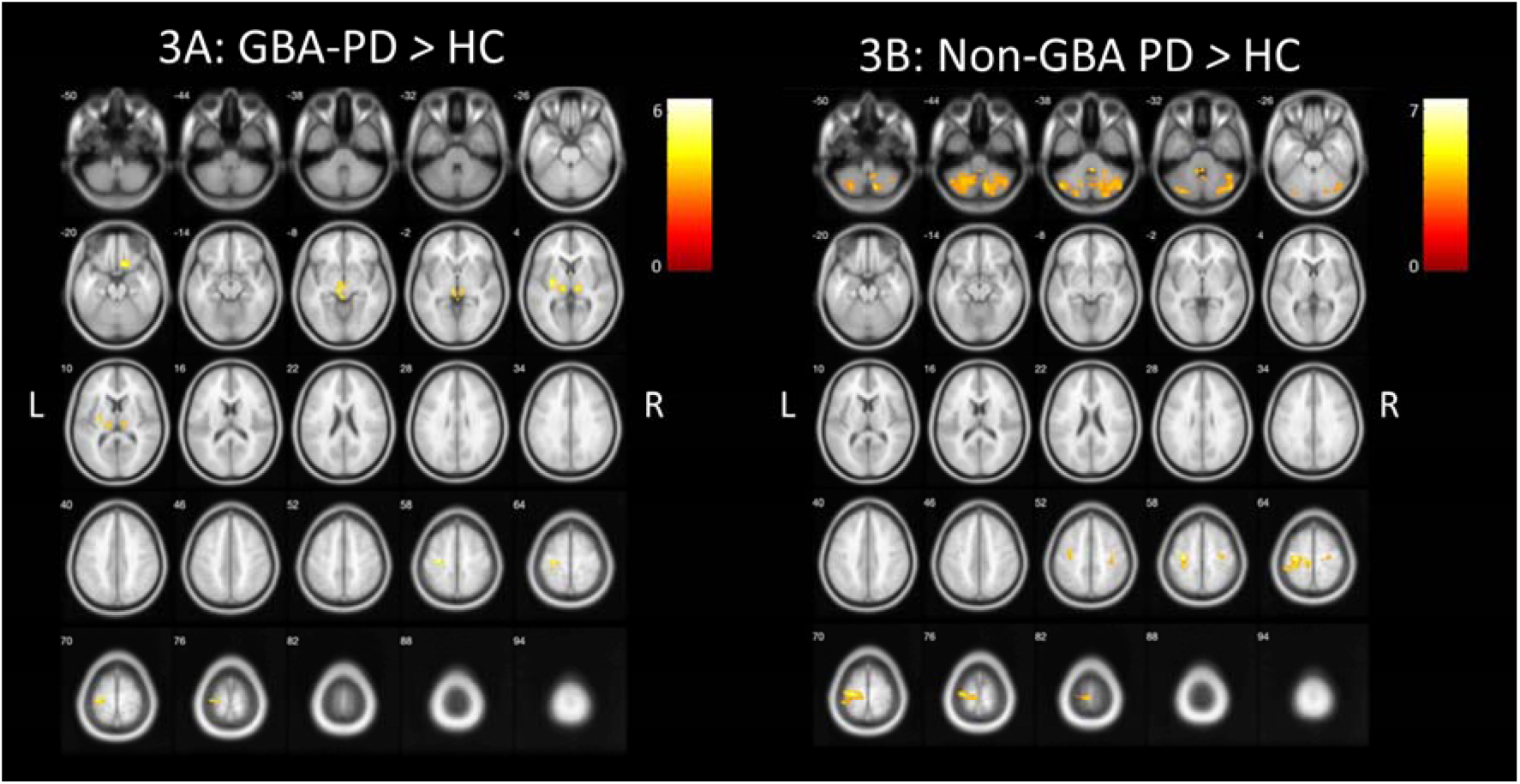
Whole brain voxel-based analyses showing significant higher VAChT binding (*P* < 0.001, uncorrected at voxel level, cluster size 100) in GBA-PD, *n* = 17 (3A) and non-GBA-PD, *n* = 17 (3B) compared to HC, controlled for age. L, left; R, right.

#### GBA-PD *vs.* non-GBA-PD groups

The results of the bidirectional whole-brain voxel-based showed no significant voxels using FDR-correction (*P* < 0.05) at the voxel level when GBA-PD was compared to non-GBA-PD group (whole, non-GBA-PD group 1 and non-GBA-PD group 2). To gain a general view of higher and lower cholinergic topography, the results are shown without FDR correction at the voxel level (*P* < 0.05).

##### GBA *vs.* non-GBA whole group

GBA-PD patients showed lower VAChT binding in the left middle and inferior occipital gyrus, left parahippocampal gyrus, inferior temporal gyrus and middle and superior frontal gyrus and right superior parietal gyrus compared to the non-GBA-PD whole group (*Fig. 4A, P < 0.05, uncorrected).* The results of higher VAChT binding in GBA-PD compared to non-GBA-PD whole group were negligible (*Fig. 4B, P < 0.05, uncorrected*).

**Figure 4:**
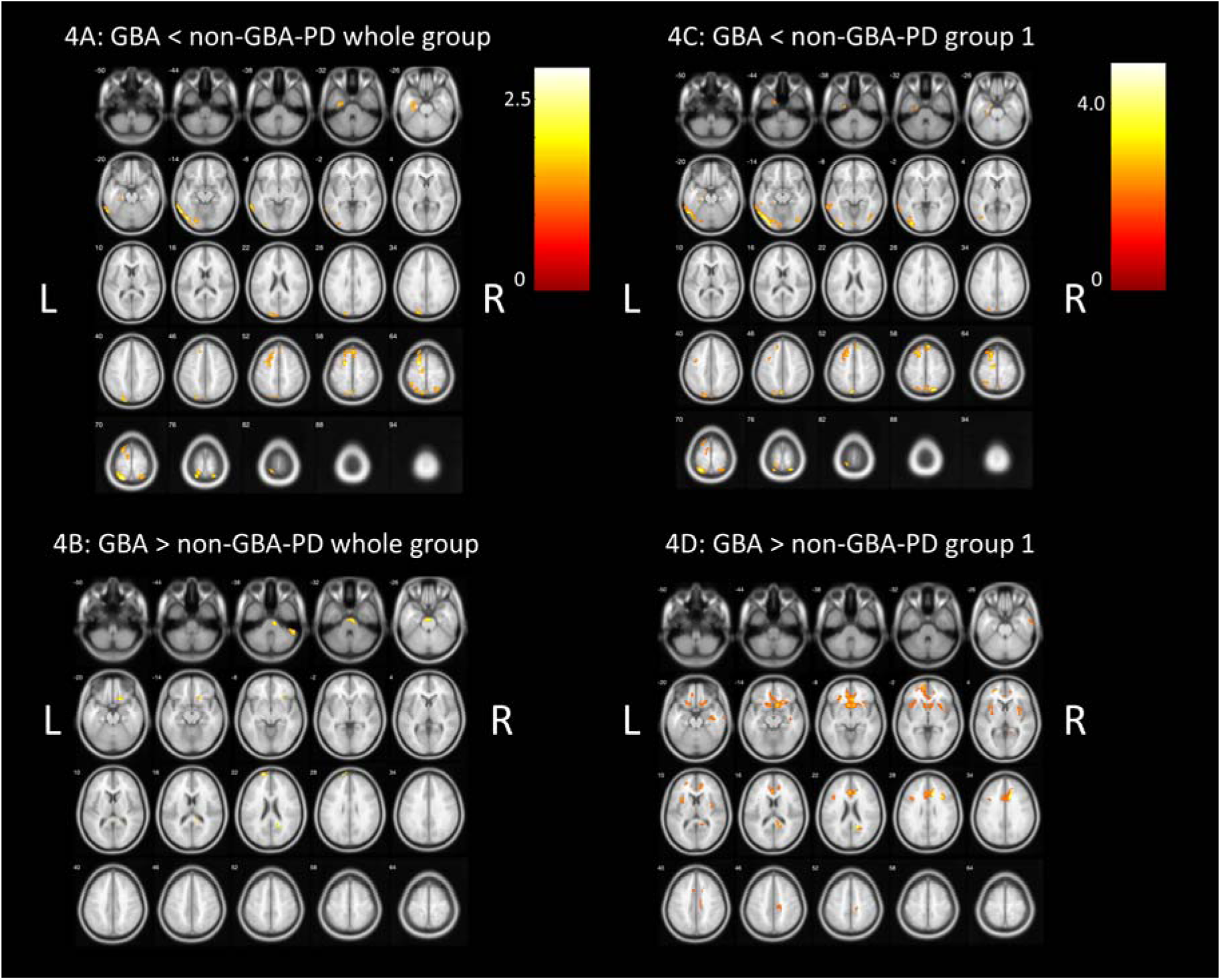
Whole brain voxel-based analyses showing significant lower (4A+C) and higher (4B+D) VAChT binding in GBA-PD, *n* = 17 compared to non-GBA-PD whole group, *n* = 106 (4A+B) and GBA-PD, *n* = 17 compared to non-GBA-PD (group 1), *n* = 17 (4C+D). *P* < 0.05, uncorrected at voxel level, cluster size 100. L, left; R, right.

##### GBA *vs*. non-GBA group 1

Lower VAChT binding in GBA-PD compared to the matched non-GBA-PD group 1 included lower binding in the left middle and inferior occipital gyrus and left middle and superior frontal gyrus (*Fig. 4C, P < 0.05, uncorrected*). GBA-PD also showed higher cholinergic innervation in the right anterior and posterior cingulate gyrus and left anterior cingulate compared to non-GBA group 1 (*Fig. 4D, P < 0.05, uncorrected*).

##### GBA *vs*. validation group (non-GBA group 2)

In the left middle and inferior occipital gyrus, inferior temporal gyrus, precuneus, superior parietal lobule, middle frontal gyrus GBA-PD, and parahippocampus gyrus (left > right) showed lower VAChT binding compared to non-GBA-PD group 2 (*Supplementary Figure. 1A, P < 0.05, uncorrected*). Reversed direction show higher VAChT binding in the pons, left superior frontal gyrus, right precuneus and right inferior frontal gyrus (Brodmann area 47) in GBA-PD (*Supplementary Figure. 1B, P < 0.05, uncorrected*).

### Volume-of-interest (VOI) analyses

VOI analyses showed no significant lower or higher VAChT binding regions between PD groups (GBA-PD *vs.* non-GBA-PD) after Bonferroni correction. Both GBA-PD and non-GBA-PD groups showed lower DVR compared to HC in occipital parietal and temporal regions (see *Table 3 for the significant VOI* and see *Supplementary Table 1 for all VOI*). However, GBA-PD had a more widespread lower [^18^F]FEOBV binding in parieto-temporal regions, including the left cuneus, entorhinal cortex, fusiform gyrus and supramarginal gyrus, when compared with HCs. Also, a significant higher VAChT binding mechanism (DVR higher than HC) emerged only in the GBA-PD group, in the bilateral medial orbitofrontal cortex and left caudal anterior cingulate cortex. All significant VOIs are shown in *Table 3*.

**Table 3:** Significant volume-of-interest (VOI) of GBA-PD, non-GBA-PD and HC subjects after Bonferroni correction.

| Volume-of-Interest | GBA-PD,<br>*n* = 17 | Non-GBA-PD<br>(group 1),<br>*n* = 17 | HC,<br>*n* = 10 | GBA-PD vs.<br>HC<br>*P*-value | Non-GBA-PD<br>(group 1) vs.<br>HC<br>*P*-value |
| --- | --- | --- | --- | --- | --- |
| Caudal anterior-cingulate<br>cortex Left (L) | 1,57±0,15 | 1,48±0,19 | 1,54±0,09 | **0,041<sup>a</sup>** | 1,000 |
| Cuneus cortex L | 0,85±0,11 | 0,88±0,1 | 1,03±0,09 | **0,022<sup>b</sup>** | 0,116 |
| Cuneus cortex Right (R) | 0,84±0,1 | 0,85±0,1 | 1,03±0,08 | 0,004 | 0,008 |
| Entorhinal cortex L | 1,12±0,09 | 1,17±0,14 | 1,33±0,1 | **0,006<sup>b</sup>** | 0,226 |
| Fusiform gyrus L | 0,99±0,09 | 1,04±0,09 | 1,15±0,05 | **0,009<sup>b</sup>** | 0,417 |
| Inferior parietal cortex L | 0,9±0,1 | 0,92±0,06 | 1,05±0,07 | 0,002 | 0,018 |
| Inferior parietal cortex R | 0,93±0,09 | 0,93±0,07 | 1,06±0,06 | 0,026 | 0,027 |
| Lateral occipital cortex L | 0,82±0,08 | 0,85±0,06 | 1±0,06 | 0,001 | 0,003 |
| Lateral occipital cortex R | 0,84±0,09 | 0,85±0,06 | 1±0,07 | 0,002 | 0,003 |
| Lateral orbito frontal cortex R | 1,37±0,07 | 1,33±0,1 | 1,27±0,05 | 0,001 | 0,028 |
| Medial orbito frontal cortex L | 1,43±0,09 | 1,38±0,12 | 1,33±0,06 | **0,004<sup>a</sup>** | 0,108 |
| Medial orbito frontal cortex R | 1,38±0,09 | 1,34±0,12 | 1,27±0,06 | **0,005<sup>a</sup>** | 0,110 |
| Middle temporal cortex L | 1,03±0,09 | 1,04±0,07 | 1,17±0,05 | 0,013 | 0,037 |
| Middle temporal cortex R | 1,06±0,1 | 1,06±0,09 | 1,18±0,04 | 0,118 | 0,074 |
| Pericalcarine cortex L | 0,91±0,15 | 0,93±0,13 | 1,16±0,1 | 0,007 | 0,018 |
| Pericalcarine cortex R | 0,91±0,12 | 0,9±0,1 | 1,15±0,11 | 0,001 | 0,001 |
| Supramarginal gyrus L | 1,02±0,12 | 1,03±0,09 | 1,19±0,13 | **0,029<sup>b</sup>** | 0,061 |
| Transverse temporal cortex L | 1,21±0,29 | 1,23±0,23 | 1,68±0,2 | 0,013 | 0,029 |
| Transverse temporal cortex R | 1,21±0,27 | 1,21±0,27 | 1,67±0,17 | 0,012 | 0,013 |

<sup>a</sup> GBA-PD showed significantly higher VACHT binding compared to healthy controls in this VOI, whereas non-GBA-PD did not.
<sup>b</sup> GBA-PD showed significant lower VACHT binding compared to healthy controls in this VOI, whereas non-GBA-PD groups 1 did not.

## Discussion

We investigated clinical features and regional cholinergic terminal changes in *de novo* Parkinson’s disease subjects with and without relevant *GBA1* mutations. To our knowledge, this is the first assessment of regional cholinergic innervation in GBA-PD. We did not find any significant differences in demographics, clinical motor or non-motor characteristics between GBA-PD and non-GBA-PD at the time of diagnosis. Although the two Parkinson’s disease groups were not distinguishable by clinical features, *GBA1* mutation carriers showed more extensive cholinergic denervation – involving more temporo-parietal regions – than non-carriers compared to healthy controls. Our results suggest that *GBA1* mutations are associated with cholinergic deficits and more widespread neurodegeneration in early disease.

*GBA1* mutation carriers were 13.8% of Parkinson’s disease patients in our cohort, consistent with recent large nationwide genetic screening of more than 3000 participants in the Netherlands.^25^ Whole-gene *GBA1* sequencing revealed that the majority (65%) of our GBA-PD patients have the E326K and T369M variants. These *GBA1* variants are the most common ones in the Netherlands.^25^ These variants are associated with Parkinson’s disease but not with Gaucher’s disease and normally present with a milder phenotype than other *GBA1* mutations associated with Parkinson’s disease.^34^ The prevalence of these variants in our cohort might explain why we did not find differences in the age of onset between Parkinson’s patients with and without *GBA1* mutations.^1, 35^ Early age of disease onset is mainly observed in carriers of severe *GBA1* variants.^10, 36^

As previously reported,^37^ GBA-PD and non-GBA-PD were clinically indistinguishable at the time of diagnosis. We did not find significant differences in UPDRS motor scores, motor phenotypes, or Hoehn and Yahr stages between *GBA1* mutant carriers and non-carriers. Previous studies also did not find significant differences in motor scores at early disease stages.^3, 37, 38^ Although higher prevalence of postural instability and gait disorders motor phenotype was documented in GBA-PD,^39^ this was not found in our GBA-PD group. These discrepancies might be due to our limited sample of *GBA1* patients, the prevalence of mild mutations, and evaluation at diagnosis. Differences in motor severity between GBA and non-GBA-PD seem to emerge during later disease stages.^5^ Longitudinal assessment of DUPARC participants will provide more information on differences in disease progression between GBA-PD and non-GBA-PD carriers.

We found comparable neuropsychological performance in GBA-PD carriers and non-GBA-PD. These findings are consistent with prior studies describing GBA-PD at early disease stages.^3, 37, 40, 41^ Previous longitudinal studies suggest divergence rate of cognitive deterioration approximately three years after diagnosis.^37, 41^

Both our voxel-wise and VOI-based analyses showed that cholinergic terminal deficits were more pronounced in GBA-PD than in non-GBA-PD. Although regional overlap exists, GBA-PD patients had more extensive deficits in parieto-temporal regions. VOI analysis results showed significantly lower [^18^F]FEOBV binding in the left cuneus, entorhinal cortex, fusiform gyrus and supramarginal gyrus in GBA-PD. This denervation topography is consistent with previous cholinergic imaging studies, which described predominantly posterior and temporal cortical binding differences between PD and HC.^16, 20, 42^ These studies generally focused on more advanced Parkinson’s disease subjects. Our results suggest that GBA-PD exhibits accelerated degeneration of especially nucleus basalis of Meynert (Ch4) projections neurons.

We previously described significantly higher [^18^F]FEOBV binding in several regions in *de novo* Parkinson’s disease subjects, suggesting compensatory upregulation of cholinergic systems at early disease phases. Higher regional [^18^F]FEOBV binding was not seen in PD-MCI subjects, suggesting that loss of cholinergic compensation plays a role in progression to PD-MCI.^18^ In the present study, we did not find significantly higher [^18^F]FEOBV binding in the GBA-PD group, when compared to controls in voxel-based analyses, with the non-GBA-PD group exhibiting higher [^18^F]FEOBV binding in some frontal and parietal regions. These results suggest that GBA-PD exhibited blunted cholinergic compensatory mechanisms at the time of diagnosis, consistent with more advanced impairment of basal forebrain cholinergic projection neurons. Additionally, the cortex of the cerebellum showed higher [^18^F]FEOBV binding in non-GBA-PD when compared to controls. Early lack of cholinergic upregulation in the cerebellum of GBA-PD might contribute to faster gait and balance disfunction.^43^

In this study we used a voxel-based as well as a VOI-based approach. Both showed widespread lower [^18^F]FEOBV binding in GBA-PD than non-GBA-PD in left parieto-temporal regions when compared to controls. The consistency of the results from both analyses supports our conclusions. VOI analyses comparing the higher binding areas of PD groups with controls did not show significant higher binding in the cerebellum and thalamus, which was expected based on the voxel-based results. It is plausible that significant higher [^18^F]FEOBV binding regions were demonstrated if the cerebellum and thalamus were divided in specific VOI regions. The interpretation of results based on combining VOI and voxel-based analysis, in which consistent or opposing results are found, demonstrates the complementary value of using both approaches.

Relevant *GBA1* mutations are related to loss of glucocerebrosidase activity and lysosomal dysfunction. Parkinson’s disease is a heterogeneous syndrome and efforts to subtype Parkinson’s disease using clinical measures are largely sterile.^44^ An alternative approach is to subtype Parkinson’s disease on the basis of pathogenic mechanisms.^45^ GBA-PD would potentially represent one such subtype. It is possible that early basal forebrain cholinergic projection system degeneration might be characteristic of a lysosomal Parkinson’s disease subtype. Application of [^18^F]FEOBV imaging to subjects with other potential forms of lysosomal Parkinson’s disease and discovery of early, widespread basal forebrain cholinergic projection system degeneration would support the existence of a lysosomal Parkinson’s disease subtype.^46^ Although no cognitive or other clinical differences were observed between the GBA-PD and non-GBA-PD groups, the distinct cholinergic topography, characterized by more widespread cholinergic denervation and limited upregulation, may be a predictor of a more aggressive clinical trajectory in GBA-PD. Longitudinal assessment is needed to explore this possibility.

As far as we know, this is the first study that assessed cholinergic system changes in GBA-PD *in vivo*. Previous studies using different imaging techniques reported reduced cortical activity and reduced cerebral blood flow in parietal and occipital cortices.^9, 47^ A recent comprehensive review on neuroimaging in GBA-PD assessed all imaging techniques used to date in cross-sectional or longitudinal studies to differentiate GBA-PD from non-GBA-PD.^48^ Nigrostriatal imaging studies reported, similar to our results, mostly no differences^47, 49, 50^; however, also contrasting findings were found varying from reduced,^9, 34^ to increased dopaminergic levels.^51^ Differences in sample size, stage of Parkinson’s disease, and methods used for genotyping in these studies were suggested as a possible explanation for contrasting results.^48^ A Fluorodeoxyglucose (FDG)-positron emission tomography (PET) study showed a more typical expression of cortical glucose metabolism – Parkinson’s disease related pattern (PDRP) - in GBA-PD compared to non-GBA-PD, even when clinical differences were not (yet) significant.^34^ The PDRP includes hypometabolism in the parieto-occipital cortex and therefore resemblances the topography of cholinergic deficit in GBA-PD we found in this study.

In this study, indications for a distinct cholinergic topography when directly compare GBA-PD to non-GBA-PD were found, however after correction for multiple comparison, significant lower or higher VAChT binding was not observed. It is likely that both the GBA-PD as well as the non-GBA groups are quite heterogenous, including Parkinson’s disease patients with mild and severe subtypes. It should be noted that we only performed whole-gene *GBA1* screening and did not screen for other Parkinson’s disease related mutations in this study. Besides *GBA1*, also other (genetic) factors are known as predictors of cognitive deterioration in Parkinson’s disease, such as variants in the apolipoprotein E, microtubule-associated protein tau and a-synuclein loci.^52–54^ Also, increased acetylcholinesterase activity has been reported in carriers of leucine repeat kinase 2 mutations with prodromal Parkinson’s disease.^15, 55^ The expected diversity of both Parkinson’s disease groups, except from having or not having a *GBA1* mutation, could be an important factor we did not found differences on clinical and cholinergic level in the direct comparison.

### Strengths and limitations

A major strength of this study is the unbiased inclusion of *de novo* Parkinson’s disease subjects. Genetic screening was performed after the collection of the clinical data and therefore, all assessors were blinded to the genetic status of the patients. Our study included an extensive and detailed subject assessment, including extensive neuropsychological test battery, clinical assessment, putaminal dopaminergic evaluation and the use of [^18^F]FEOBV PET. The extensive cognitive assessment in this study allowed for type II classification of PD-MCI,^27^ and domain specific cognitive performance. Previous *in vivo* imaging studies of brain cholinergic systems in Parkinson’s disease mostly relied on PET acetylcholinesterase substrate tracers, which are indirect markers of cholinergic terminal integrity because acetylcholinesterase may have both pre-and post-synaptic expressions and its activity may be regulated in disease states.^56^ The big advantage of [^18^F]FEOBV PET imaging is the strict correlation with presynaptic cholinergic terminals.^20, 57^ A final strength is the use of a validation cohort comparing GBA-PD with a second matched non-GBA-PD group to validate our results.

A limitation of this study is the relatively small sample size of the GBA-PD group. Due to the heterogeneity within Parkinson’s patients, significant differences might have been missed, due to limited power. Future studies with larger sample sizes would allow for more detailed stratification, for example, by mutation or sex, which will improve our understanding of the cholinergic system pathologies of GBA-PD.

Furthermore, the HC group had a significantly lower age than both PD groups. An important role of age in the relationship between cognition and cholinergic innervation has been demonstrated.^58^ Although the analyses were corrected for age, a possible role of the age difference cannot be ruled out.

The cross-sectional design does not allow longitudinal assessment of cholinergic innervation changes. Further research should enhance our understanding of the role of cholinergic dysfunction in GBA-PD with longitudinal study. Clarification of the relationship between cholinergic deficits and characterisation of GBA-PD may be useful for defining a Parkinson’s disease subtype. Longitudinal data on the cholinergic profile of our GBA-PD patients can be expected in the near future.

## Conclusion

The results of this study provide the first data on the cholinergic systems in GBA-PD subjects. This study demonstrates evidence of deficits in cortical cholinergic innervation in *de novo* PD, with and without *GBA1* mutations. At the time of diagnosis, despite a similar clinical profile, *GBA1* mutation carriers showed more extensive cholinergic denervation than non-GBA-PD. This distinct cholinergic deficit topography suggests more advanced cholinergic degeneration in GBA-PD, potentially contributing to the faster disease progression of this subset of Parkinson’s disease subjects.

### Abbreviations

[^18^F]FDOPA: 3,4-dihydroxy-6-18F-fluoro-I-phenylalanine;
[^18^F]FEOBV: [^18^F]Fluoroethoxybenzovesamicol;
DUPARC: Dutch Parkinson Cohort;
DVR: Distribution Volume Ratios;
FDR: False Discovery Rate;
GBA-PD: Parkinson’s disease patients who carry *GBA1* mutations;
HC: Healthy controls;
Non-GBA-PD: Parkinson’s disease patients without *GBA1* mutations;
PD-MCI: Parkinson’s disease Mild Cognitive Impairment;
VOI: Volume-of-interest;
UPDRS: Unified Parkinson’s disease Rating Scale;
VAChT: Vesicular Acetylcholine Transporter

## Data Availability

All data produced in the present study are available upon reasonable request to the authors.

## Acknowledgements

We thank all patients, caregivers, health-care professionals, and students who have contributed to and collaborated in this project. The inclusion of patients was established with help of the collaborative Parkinson Platform Northern Netherlands.

## Supplementary material

**Supplementary table 1.** Volumes of Interest (VOI) subject specific.

|  |  |
| --- | --- |
| **Cortical-cerebellar VOIs** | Superior temporal gyrus |
| Caudal anterior-cingulate cortex | Supramarginal gyrus |
| Caudal middle frontal gyrus | Temporal pole |
| Cerebellum cortex | Transverse temporal cortex |
| Cuneus cortex | **Subcortical VOIs** |
| Entorhinal cortex | Accumbens Area |
| Frontal Pole | Amygdala |
| Fusiform gyrus | Hippocampus |
| Inferior parietal cortex | Caudate Nucleus |
| Inferior temporal gyrus | Putamen |
| Insula | Thalamus |
| Lateral occipital cortex | Globus Pallidum |
| Lateral orbital frontal cortex | Brainstem |
| Lingual gyrus |  |
| Medial orbital frontal cortex |  |
| Middle temporal gyrus |  |
| Parahippocampal gyrus |  |
| Paracentral lobule |  |
| Inferior Frontal Gyrus Pars opercularis |  |
| Inferior Frontal Gyrus Pars orbitalis |  |
| Inferior Frontal Gyrus Pars triangularis |  |
| Pericalcarine cortex |  |
| Postcentral gyrus |  |
| Posterior-cingulate cortex |  |
| Precentral gyrus |  |
| Precuneus cortex |  |
| Rostral anterior cingulate cortex |  |
| Rostral middle frontal gyrus |  |
| Superior frontal gyrus |  |
| Superior parietal cortex |  |

**Supplementary table 2:** Overview of *GBA1* variants.

| DNA sequencing of complete human *GBA1*-gene, using Illumina NovaSeq6000 sequencing.<br>The *GBA1* allelic names are given, excluding the 39 amino acid signalling peptide. |  |  |  |
| --- | --- | --- | --- |
| Number of subjects | *GBA1* mutations (allelic name) | Type of mutation | Position Chr1 |
| 5 | p.T369M | missense variant & splice region variant | chr1:155206037:A |
| 6 | p.E326K | missense variant | chr1:155206167:T |
| 2 | p.E326K;p.D140H | missense variant;missense variant | chr1:155206167:T;chr1:155208361:G |
| 1 | p.V460=;p.A456P | synonymous variant;missense variant | chr1:155204994:G;chr1:155205008:G |
| 1 | p.L444P;p.V460=;p.A456P | missense variant;synonymous variant;missense variant | chr1:155205043:G;chr1:155204994:G;chr1:155205008:G |
| 1 | p.N370S;p.R329C;p.K346E | missense variant & splice region variant;missense variant;missense variant | chr1:155205634:C;chr1:155206158:A;chr1:155206107:C |
| 1 | p.F213V | missense variant | chr1:155207932:C |

**Supplementary table 3:** Demographics and Clinical Characteristics of the study participants, comparing HC, GBA-PD and non-GBA-PD.

|  | HC<br>(n = 10) | GBA-PD<br>(n = 17) | non-GBA-PD<br>(n = 17) | P-value |
| --- | --- | --- | --- | --- |
| Age y<sup>a</sup> | 54.60 (6.02) | 65.18 (10.27) | 65.35 (9.94) | **0.011<sup>b</sup>** |
| Gender, male n (% male) | 5 (50.0%) | 10 (58.8%) | 10 (58.8%) | 0.892 |
| Educational level<sup>c</sup> | 5.0 [5.0-6.25] | 5.0 [4.0-6.0] | 5.0 [4.0-5.0] | 0.167 |
| Putaminal dopaminergic<br>striatal-to-occipital ratio,<br>most affected side |  | 1.79[1.67-1.95] | 1.93[1.80-1.99]* | 0.087 |
| *Motor symptoms* |  |  |  |  |
| Motor symptom duration |  | 24.00 [8.25-30.00]** | 12.00 [6.50-22.00] <sup>‡</sup> | 0.151 |
| UPDRS-III, total score |  | 29.24 (13.42) | 32.76 (9.86) | 0.325 |
| Motor phenotype,<br>n TD/PIGD/indeterminate |  | 6/7/4<br>(35%/41%/24%) | 8/7/2<br>(47%/41%/12%) | 0.618 |
| Hoehn and Yahr stage |  | 2.00 [1.00-2.00] | 2.00 [2.00-2.00] | 0.339 |
| *Non-motor symptoms* |  |  |  |  |
| MoCA, total score | 28.4 (0.84) | 25.76 (2.82) | 25.00 (3.46) | **0.014<sup>d</sup>** |
| PD-MCI |  | 5 (29.4%) | 5 (29.4%) | 1.000 |
| Z-score memory |  | -0.06(0.73) | -0.28 (0.75) | 0.393 |
| Z-score attention |  | -0.72 (0.66) | -0.48 (0.84) | 0.362 |
| Z-score executive function |  | -0.38 (0.72) | -0.22 (0.83) | 0.552 |
| Z-score language |  | -0.07 (0.69) | -0.26 (0.98) | 0.560 |
| Z-score visuospatial function |  | -0.11 (0.97) | -0.17 (0.97) | 0.859 |
| NMS-Quest, total score |  | 4.0 [3.00-7.75]<sup>§</sup> | 6.00 [4.00-8.50] | 0.337 |
| HADS anxiety, total score |  | 4.06 (3.07) | 4.29 (2.47) | 0.335 |
| HADS depression, total score |  | 2.00 [2.00-4.00] | 5.00 [1.00-7.00] | 0.245 |
| RBD Quest, total score |  | 3.00 [1-4.50] | 2.00 [1.00-6.00] | 0.819 |
| Sniffin' sticks, total score |  | 5.06 (2.33) | 5.64 (2.56) | 0.356 |

Abbreviations: UPDRS-III, Movement Disorders Society Unified Parkinson's Disease Rating Scale part III; TD, Tremor Dominant; PIGD, Postural Instability and Gait Difficulty; MoCA, Montreal Cognitive Assessment; PD-MCI, Parkinson's Disease Mild Cognitive Impairment; NMS-Quest, Non-Motor Symptoms Questionnaire; HADS, Hospital Anxiety and Depression Scale; RBD Quest, REM Sleep Behaviour Disorder Screening Questionnaire; Sniffin' sticks, Burghart Sniffin' Sticks 12 Tests
a. mean (standard deviation); b. post-hoc analysis: HC < GBA-PD, HC < non-GBA-PD; c. median [Q1- Q3]; d. post-hoc analysis: HC > GBA-PD, HC > non-GBA-PD
\* missing n = 1, \*\* missing n = 1, <sup>‡</sup> missing n = 4, <sup>§</sup> missing n = 1

**Supplementary table 4:**
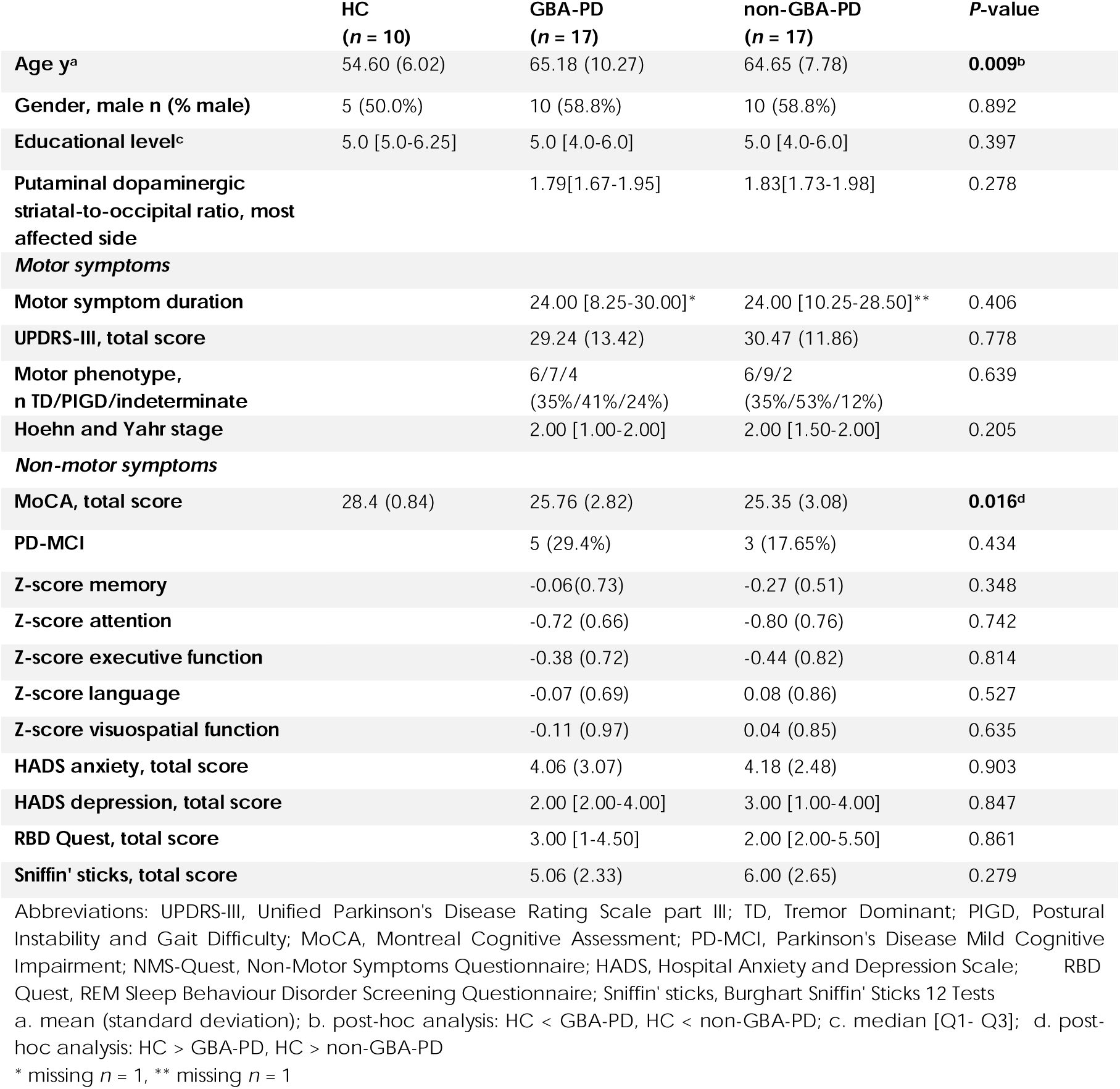
Demographics and Clinical Characteristics of the study participants, comparing HC, GBA-PD and second non-GBA-PD group 2.

|  | HC<br>(n = 10) | GBA-PD<br>(n = 17) | non-GBA-PD<br>(n = 17) | P-value |
| --- | --- | --- | --- | --- |
| Age y<sup>a</sup> | 54.60 (6.02) | 65.18 (10.27) | 64.65 (7.78) | **0.009<sup>b</sup>** |
| Gender, male n (% male) | 5 (50.0%) | 10 (58.8%) | 10 (58.8%) | 0.892 |
| Educational level<sup>c</sup> | 5.0 [5.0-6.25] | 5.0 [4.0-6.0] | 5.0 [4.0-6.0] | 0.397 |
| Putaminal dopaminergic<br>striatal-to-occipital ratio, most<br>affected side |  | 1.79[1.67-1.95] | 1.83[1.73-1.98] | 0.278 |
| *Motor symptoms* |  |  |  |  |
| Motor symptom duration |  | 24.00 [8.25-30.00]* | 24.00 [10.25-28.50]** | 0.406 |
| UPDRS-III, total score |  | 29.24 (13.42) | 30.47 (11.86) | 0.778 |
| Motor phenotype,<br>n TD/PIGD/indeterminate |  | 6/7/4<br>(35%/41%/24%) | 6/9/2<br>(35%/53%/12%) | 0.639 |
| Hoehn and Yahr stage |  | 2.00 [1.00-2.00] | 2.00 [1.50-2.00] | 0.205 |
| *Non-motor symptoms* |  |  |  |  |
| MoCA, total score | 28.4 (0.84) | 25.76 (2.82) | 25.35 (3.08) | **0.016<sup>d</sup>** |
| PD-MCI |  | 5 (29.4%) | 3 (17.65%) | 0.434 |
| Z-score memory |  | -0.06(0.73) | -0.27 (0.51) | 0.348 |
| Z-score attention |  | -0.72 (0.66) | -0.80 (0.76) | 0.742 |
| Z-score executive function |  | -0.38 (0.72) | -0.44 (0.82) | 0.814 |
| Z-score language |  | -0.07 (0.69) | 0.08 (0.86) | 0.527 |
| Z-score visuospatial function |  | -0.11 (0.97) | 0.04 (0.85) | 0.635 |
| HADS anxiety, total score |  | 4.06 (3.07) | 4.18 (2.48) | 0.903 |
| HADS depression, total score |  | 2.00 [2.00-4.00] | 3.00 [1.00-4.00] | 0.847 |
| RBD Quest, total score |  | 3.00 [1-4.50] | 2.00 [2.00-5.50] | 0.861 |
| Sniffin' sticks, total score |  | 5.06 (2.33) | 6.00 (2.65) | 0.279 |

Abbreviations: UPDRS-III, Unified Parkinson's Disease Rating Scale part III; TD, Tremor Dominant; PI GD, Postural Instability and Gait Difficulty; MoCA, Montreal Cognitive Assessment; PD-MCI, Parkinson's Disease Mild Cognitive Impairment; NMS-Quest, Non-Motor Symptoms Questionnaire; HADS, Hospital Anxiety and Depression Scale; RBD Quest, REM Sleep Behaviour Disorder Screening Questionnaire; Sniffin' sticks, Burghart Sniffin' Sticks 12 Tests
a. mean (standard deviation); b. post-hoc analysis: HC < GBA-PD, HC < non-GBA-PD; c. median [Q1- Q3]; d. post-hoc analysis: HC > GBA-PD, HC > non-GBA-PD
\* missing n = 1, \*\* missing n = 1

**Supplementary figure 1:**
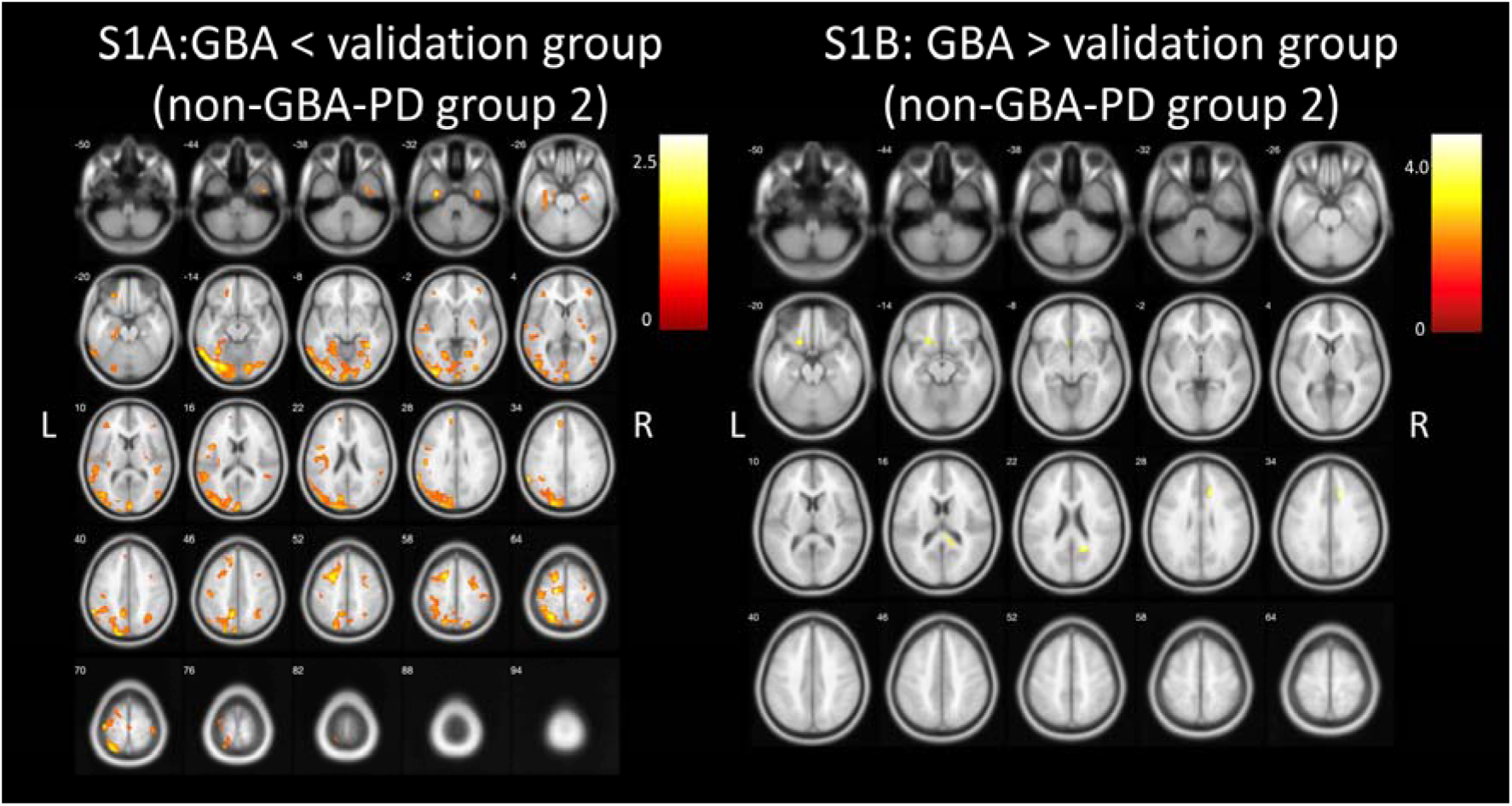
Whole brain voxel-based analyses showing significantly lower (1A) and higher (1B) VAChT binding (*P* < 0.05) in GBA-PD, *n* = 17 compared to non-GBA-PD group 2, *n* = 17 uncorrected at voxel level, cluster size 100. L, left; R, right.

